# Factors Influencing Health Insurance Enrollment and Its Impact on Outpatient Service Utilization in Saudi Arabia: Insights from the National Saudi Family Health Survey

**DOI:** 10.1101/2024.10.17.24315658

**Authors:** Khaled Shaeel Althabaiti, Monica Hunsberger, Jahangir Khan, Sayem Ahmed

## Abstract

The Kingdom of Saudi Arabia (KSA) recently launched a reform plan for its health care system in 2021 driven by its Vision 2030 initiative. This vision aims to reduce dependence on government resources by transitioning to the national health insurance model and the Cooperative Health Insurance program, especially for the immigrant population. This reform may impact the utilization of health services by citizenship and insurance status. The current study aims to identify factors influencing health insurance enrollment and its impact on outpatient service utilization in the Kingdom of Saudi Arabia. This study used data from the 2018 Saudi Family Health Survey (FHS). The survey covers a nationally representative sample from KSA (n=8,274), which contains questions that obtain information about the health insurance enrollment, health care utilization, chronic disease condition, and health status of the respondents. We conducted a bivariate analysis using a chi-square test and an independent-sample t-test to examine the significance of differences between groups (by nationality and insurance status). We employed multiple binary logistic regression models to measure the association between health insurance enrollment and the demographic and socioeconomic characteristics of the respondents. Further, the multiple Poisson regression model was used to estimate the effect of health insurance status on the utilization of outpatient care. Most of the respondents were Saudis (76.8%), and the number of males (54.9%) respondents were higher than the females. Around 26.2% of the total respondents were insured and the proportion of insured was significantly higher among non-Saudis (72.8%) compared to Saudis (12.1%). The logistic regression showed that individuals with a high monthly income, non-Saudi, males, being married, high level of education, and perceived good health were associated with health insurance enrollment. We found health insurance enrollment was associated with lower utilization of outpatient services (co-efficient -0.107; P<0.001). Other factors increasing utilization of outpatient services were being female, having a high monthly income, being never married, having chronic diseases, and the perception of bad health. Significant determinants of health insurance enrollment were being non-Saudi, males, having a high income, higher education level, and perceived good health status. However, health insurance was associated with lower utilization of outpatient services. The results of the current study should be taken into consideration when planning for the implementation and monitoring reform of the health system in Saudi Arabia.

## INTRODUCTION

The Kingdom of Saudi Arabia provides health care services free of charge to its citizens (1). Non-Saudis who are working in the governmental sector have similar access to health care. However, the government realized that this model is unsustainable in the long run, especially with an increased number of expatriates who make up a large proportion of the population (41.6% in the year 2022), and with the increased demand for health care services (2,3). Consequently, there was an inevitable need for health care reform to ensure the continuity of providing adequate health care services (3). The decision-makers decided to transition towards the National Health Insurance model. The first step was taken by the passage of the Council of Cooperative Health Insurance Law in 1999, which established a mandatory health insurance scheme Cooperative Health Insurance (CHI) in 2002 for private sector employees, who were mostly expatriates (4). The scheme aimed to relieve the financial burden on the public sector by forcing private sector employers who form 67.9% of the workforce in Saudi Arabia to cover health care costs for their Saudi employees (22.3%) and non-Saudis (77.7%). This scheme will push the private sector employees to private health care providers to get their health services (5,6).By definition, health care services are the medical services provided by health care professionals to treat, protect or promote the health and well-being of the people; it includes hospital care (emergency care, outpatient clinics, admissions, diagnostic measures), primary health care services, physicians and clinical services, dental services; besides home health care, other residential health services, prescribing drugs, durable and nondurable medical products (7).

The Ministry of Health (MOH) in Saudi Arabia is responsible for public health care services and other governmental sectors, such as the National Guard and educational health facilities, in addition to the private sector which also provides health care services (8). According to the reports published by the MOH in 2022, the number of visits to the health care facilities in the public sector (hospitals and primary health care centers) accounted for 78.6 million (61%), while it reached 20.4 million (16%) in other governmental sectors and 29.5 million (23%) in the private sector (8). The number of visits has been accepted as a quantification of the utilization of health care services (9). According to the theoretical model of health service use provided by Andersen in 1973, the factors determining the extent of health care utilization can be grouped under three main categories; “predisposing factors” represented by the demographic characteristics such as age and gender and attitude of the individual; the “enabling factors” described by the availability and accessibility of health care providers, and the “needs” which are the Symptoms or health issues prompting individuals to seek medical attention (10). In the past few decades, the Gulf Cooperation Countries (GCC), including Saudi Arabia, have witnessed rapid growth in health care investments, in the form of health care facilities, medical complexes (hospitals providing secondary and tertiary care), and medical cities or groups of hospitals providing comprehensive services (11). Meanwhile, the government of Saudi Arabia introduced the Cooperative Health Insurance Law that demands insurance coverage for all employees in private companies. The strategy aimed to ease the challenge experienced by offering free health care services to the residents of the country. This strategy is offering medical coverage to more than eleven million insured people in Saudi Arabia through 27 insurance companies (12).

In Saudi Arabia, the health insurance was started in 2006 and its implementation is separated into three phases as part of the country’s broader health care reforms under Vision 2030. The first phase included coverage for employees of large companies employing 500 or more individuals, gradually extending to smaller businesses, and eventually covering all employees including Saudis and non-Saudis in the private sector(13). The second phase of the Cooperative Health Insurance scheme is intended to cover those working in the government sector. The third phase is intended to be extended to cover short-term for non-Saudi workers and visitors (e.g., tourists) this phase was still pending (14). Currently, the implementation of the health insurance is in the second phase.

There are three types of health insurance in Saudi Arabia: firstly, the government-funded health care which covers the health services in public facilities provided to the Saudi population and non-Saudis working in the government sector (15). Secondly, is employer-sponsored health insurance where the employer in the private sector takes the responsibility of payment for enrollment and renewal of insurance for the employees of both citizens and expatriates (16). According to the General Authority for Statistics in 2023, this type of insurance covered 37.5% of total the adult population aged 15 and above (20.5% for citizens and 53.6% for expatriates) (17). Thirdly, private health insurance which is available to citizens and expatriates that are paid by individuals to extend their health services benefits. In addition to international students, tourists and Pilgrims must be covered by health insurance before the entrance to the country (18).

Previous research in Saudi Arabia showed that individuals with health insurance had higher utilization of health services than the uninsured ones, especially routine check-up services (19), periodic preventive services (12), inpatient and outpatient services, visits to the emergency room (ER) (20) and dispensing medicines (21). Alrabiah and colleagues found that health insurance increased adherence to treatment and a positive perception of the quality of received services (22). Moreover, research indicated that insured individuals utilize outpatient services more frequently due to easier accessibility and the availability of a wide range of medical services (23). Despite the wide coverage of the effects of health insurance on the utilization of health care services, the literature does not give adequate attention to important characteristics of respondents that may affect the association between insurance enrollment on health-seeking behavior (19). For example, in Saudi Arabia, 41.6 % of the population are non-Saudi skilled and unskilled workers and their dependents (2) and existing studies lack exploration into the discrepancies between these two groups regarding enrollment in health insurance and utilization of health care services. This affects the understanding of possible differences in access to health care services, which can offer an exciting subject for research and investigation. Further, research study including more unexplored factors can provide a comprehensive understanding of the effect of health insurance on the utilization of health services by various demographic groups (19). By informing policy development, such insights can lead to targeted interventions aimed at improving health or health insurance literacy and increasing insurance coverage (24) and in the long term this may enhance health outcomes. In light of ongoing health care system reforms in Saudi Arabia (25), research is needed to pinpoint areas that require improvement and guide the development of more effective policies to enhance health care utilization and overall health outcomes in the country. Thus, the current study aims to identify the factors of health insurance enrollment and estimate the effect of health insurance on the utilization of outpatient services emphasizing the discrepancies between Saudi and non-Saudi populations.

This research is driven by the need to understand the relationship between health insurance enrollment and outpatient service utilization in Saudi Arabia, within the recent health care reforms, focusing on shifting from government-funded health care to health insurance systems, primarily targeting expatriates. This transition addresses the rising demand for medical services due to population growth and a significant expatriate community and should focus on health insurance literacy among workers and individuals. Understanding the factors influencing health insurance enrollment is crucial for policymakers to design effective interventions that promote increased insurance coverage and utilization of outpatient services.

## MATERIAL AND METHODS

This study used the data from the 2018 Saudi Family Health Survey (FHS). The FHS is a household survey classified under education and health statistics conducted by the General Authority for Statistics (GASTAT). This survey consists of 12,827 samples in total collected from all the 13 administrative areas in Saudi Arabia. The survey used a pre-designed validated questionnaire that had been adopted form the World Health Organization (WHO)’ Health Survey (WHS).

The survey contains questions about household’s demographic characteristics of the interviewed individuals (age, sex, nationality, education level, marital status, number of family members, monthly income, employment status and residence), health status (self-rated health condition and any chronic diseases) and the health insurance status (whether insured or not). A representative sample for the whole population in Saudi Arabia was interviewed by trained personnel using the pre-designed questionnaire (26). The data from the survey was requested officially from the sponsor of the data in the government of Saudi Arabia, represented by GASTAT. The authority provided data in Excel format that included the relevant variables for the current study. The data were translated into English and coded to facilitate statistical analysis using the SPSS ver. 26 program.

## DATA ANALYSIS

Data were summarized as frequency distribution, mean and standard deviation. Chi-squared test was carried out to test the association between the study participant’s characteristics and health insurance enrollment. An Independent sample t-test was used to examine the significance of the difference in the number of visits to health care facilities between the insured and uninsured groups. We included only outpatient care utilization in this analysis due to the availability of this in the data. Binary logistic regression was conducted to find out the predictors of health insurance enrollment. Poisson regression was employed to determine the factor associated with health care service utilization. The variables which showed significant association with the outcome variable (utilization of health services in terms of number of visits) were included in the regression analysis. Multicollinearity was assessed for the included independent variables, and the Variance Inflation Factor (VIF) values were found to be less than 5, indicating that multicollinearity is not present significance level 5% (P value<0.05) was considered for statistical tests.

## ETHICAL CLEARANCE

This study was based on the use of secondary data from the FHS, which was conducted, commissioned, funded, and managed in 2018 by GASTAT, who was in charge of all ethical procedures.

## RESULTS

Table 1 describes the demographic characteristics of the respondents, categorized according to nationality. Out of all respondents (n=8,276), 76.8% were Saudis. The proportion of the male population was higher among the non-Saudis (69.1%) than Saudis (50.6%). The highest proportion (55.80%) of the respondents were from the 26-59 age group. The respondents from this age group were 78.8% among non-Saudis and 58.7% among Saudis. Only 22.3% of total respondents had higher education, either a university qualification or a postgraduate degree. Approximately 23.3% of Saudis and 19.1% of non-Saudis had such educational qualifications.

**Table 1:** Demographic characteristics of the study group (n=8276)

| Characteristics | Total |  | Saudi |  | Non- Saudi |  | P<sup>a</sup> |
| --- | --- | --- | --- | --- | --- | --- | --- |
|  | N<br>(8,276) | % | N(6,351) | % | N<br>(1,925) | % |  |
| Sex |  |  |  |  |  |  | <0.001** |
| Male | 4,545 | 54.90% | 3,215 | 50.60% | 1,330 | 69.10% |  |
| Female | 3,731 | 45.10% | 3,136 | 49.40% | 595 | 30.90% |  |
| **Age** |  |  |  |  |  |  |  |
| 18-25 years | 1,912 | 23.10% | 1593 | 25.10% | 319 | 16.50% | <0.001** |
| 26-59 years | 5,248 | 63.40% | 3732 | 58.70% | 1,516 | 78.80% |  |
| >60 years | 1,116 | 13.50% | 1,026 | 16.20% | 90 | 4.70% |  |
| **Family monthly income (Saudi Riyal):** |  |  |  |  |  |  |  |
| <5,000 | 2,580 | 31.20% | 1,433 | 22.60% | 1,147 | 59.60% | <0.001** |
| 5000-<10000 | 2,901 | 35.10% | 2,415 | 38.00% | 486 | 25.20% |  |
| 10000-<15000 | 1,416 | 17.10% | 1,280 | 20.20% | 136 | 7.10% |  |
| 15000-<20000 | 681 | 8.20% | 612 | 9.60% | 69 | 3.60% |  |
| 20000+ | 698 | 8.40% | 611 | 9.60% | 87 | 4.50% |  |
| **Education level** |  |  |  |  |  |  |  |
| Below primary school | 1,401 | 16.90% | 1,052 | 16.60% | 349 | 18.10% | <0.001** |
| Primary school | 753 | 9.10% | 495 | 7.80% | 258 | 13.40% |  |
| Intermediate school | 1,008 | 12.20% | 651 | 10.30% | 407 | 18.50% |  |
| Secondary school | 3,270 | 39.50% | 2,675 | 42.10% | 595 | 30.90% |  |
| Higher education | 1,844 | 22.30% | 1,478 | 23.30% | 366 | 19.10% |  |
| **Marital status** |  |  |  |  |  |  |  |
| Married | 5,361 | 64.80% | 3,948 | 62.20% | 1,413 | 73.40% | <0.001** |
| Never married | 2,463 | 29.70% | 1,986 | 31.30% | 470 | 24.40% |  |
| Divorced& Widowed | 459 | 5.50% | 417 | 6.60% | 42 | 2.20% |  |
| **Employment** |  |  |  |  |  |  |  |
| Employed | 3,969 | 48.00% | 2,507 | 39.50% | 1,462 | 75.90% | <0.001** |
| Unemployed | 1,067 | 12.90% | 999 | 15.70% | 68 | 3.60% |  |
| Housewife | 2,493 | 30.10% | 2,104 | 33.10% | 389 | 20.20% |  |
| Retired | 747 | 9.10% | 741 | 11.70% | 6 | 0.30% |  |
| **Insurance** |  |  |  |  |  |  | <0.001** |
| Yes | 2,170 | 26.20% | 768 | 12.10% | 1402 | 72.80% |  |
| No | 6,106 | 73.80% | 5,583 | 87.90% | 523 | 27.20% |  |
| **Self-rating of health status:** |  |  |  |  |  |  | <0.001** |
| Very good | 5,933 | 71.70% | 4,354 | 68.50% | 1,579 | 82.00% |  |
| Good | 1,504 | 18.20% | 1,219 | 19.20% | 285 | 14.80% |  |
| Fair | 642 | 7.80% | 596 | 9.40% | 46 | 2.40% |  |
| Bad | 197 | 2.40% | 182 | 2.90% | 15 | 0.80% |  |
| **Have chronic disease** |  |  |  |  |  |  | <0.001** |
| Yes | 2,108 | 25.50% | 1,830 | 28.80% | 274 | 14.40% |  |
| No | 6,168 | 74.50% | 4,521 | 71.20% | 1,647 | 85.60% |  |
| **Type of chronic disease (n=2,108):** |  |  |  |  |  |  | 0.046** |
| Diabetes | 702 | 33.30% | 600 | 32.80% | 102 | 36.70% |  |
| Hypertension | 630 | 29.90% | 557 | 30.40% | 73 | 26.20% |  |
| Musculoskeletal | 124 | 5.80% | 113 | 6.20% | 11 | 4.00% |  |
| Hypercholesterolemia | 100 | 4.70% | 81 | 4.40% | 19 | 6.80% |  |
| Cardiovascular | 97 | 4.60% | 88 | 4.80% | 9 | 3.20% |  |
| GIT | 67 | 3.10% | 64 | 3.50% | 3 | 1.10% |  |
| Bronchial asthma | 60 | 2.80% | 53 | 2.80% | 7 | 2.50% |  |
| Others | 328 | 15.80% | 274 | 15.10% | 54 | 19.50% |  |
| **Number of health care outpatient visits in past 12 month:** |  |  |  |  |  |  |  |
| 0 | 2,122 | 25.60% | 1,214 | 19.20% | 908 | 47.30% | <0.001** |
| 1 | 2,446 | 29.60% | 1,883 | 29.60% | 563 | 29.20% |  |
| 2 | 1,959 | 23.70% | 1,709 | 26.90% | 250 | 13.00% |  |
| 3 | 993 | 12.00% | 847 | 13.30% | 146 | 7.50% |  |
| 4 | 383 | 4.60% | 360 | 5.70% | 23 | 1.20% |  |
| 5+ | 373 | 4.50% | 338 | 5.30% | 35 | 1.80% |  |
| **Number of health care visits: Median (IQR)** |  |  | 2.00;(1.00-2.00) |  | 1.00;(0-1.00) |  |  |

<sup>a</sup> Chi-squared test

Almost half of the respondents (48.0%) were employed, with a higher percentage among non-Saudis (75.9%) than Saudis (39.5%). Regarding the monthly income, a higher proportion of non-Saudis (59.6%) than Saudis (22.6%) had the lowest monthly income (<5,000 SR). One-half (64.8%) of the respondents were married, with a higher percentage of non-Saudis (73.4%) than Saudis (62.2%). One-quarter (26.2%) had health insurance and the health insurance enrollment was higher among non-Saudis (72.8%) compared to Saudi (12.1%). The majority of the Saudis (87.7%) and non-Saudis (96.8%) perceived their health status as good or very good. Chronic diseases (mostly diabetes and hypertension) were reported by 28.8% of the Saudis and 14.4% among non-Saudis. Among the Saudi respondents (48.8%) and non-Saudi respondents (76.5%) made at least one health care visit during the past 12 months. The average number of visits among Saudis were two visits with IQR=1.00-2.00, and in non-Saudis were one visit with IQR= (0.00-1.00).

Table 2 presents the health insurance enrollment based on the characteristics of the respondents. The health insurance enrollment was significantly associated with the sex of the respondents, with a higher rate of enrollment among male (32.5%) respondents compared to females (18.5%). A similar association between sex and health insurance enrollment was observed among respondents from the non-Saudi and Saudi groups. The health insurance enrollment was higher in those who aged 26-59 years (31.4%). The employed had a higher health insurance enrollment rate than the unemployed (36.4% vs 12.4%). It was observed that the percentage of the insured was significantly higher among those with lower monthly income <5,000 (36.5%). Among Saudi and non-Saudi, the highest percentage of insurance enrollment was for the income group 5000-<10000 and 15000-<20000 respectively. Marital status, education level and chronic disease were significantly associated with the health insurance enrollment.

**Table 2.** Comparing the health insurance enrollment rate of the Saudi and non-Saudi populations according to their characteristics.

| Characteristics | INSURED-Saudi |  |  | INSURED-non-Saudi |  |  | INSURED-all |  |  |
| --- | --- | --- | --- | --- | --- | --- | --- | --- | --- |
|  | N<br>(6,351) | % | P-value* | N<br>(1,925) | % | P-value* | N<br>(8,276) | % | P-value* |
| **Sex** |  |  |  |  |  |  |  |  |  |
| Male | 3,215 | 13.3 | 0.004 | 1,330 | 79.2 | <0.001 | 4,545 | 32.5 | <0.001 |
| Female | 3,136 | 10.9 |  | 595 | 58.7 |  | 3,731 | 18.5 |  |
| **Age** |  |  |  |  |  |  |  |  |  |
| 18-25 years | 1,593 | 9.5 | <0.001 | 319 | 65.5 | 0.002 | 1,912 | 18.8 | <0.001 |
| 26-59 years | 3,732 | 13.6 |  | 1,516 | 75.2 |  | 5,248 | 31.4 |  |
| >60 years | 1,026 | 10.6 |  | 90 | 58.9 |  | 1,116 | 14.5 |  |
| **Education** |  |  |  |  |  |  |  |  |  |
| Below primary school | 1,052 | 5.8 | <0.001 | 349 | 58.5 | <0.001 | 1,401 | 18.9 | <0.001 |
| Primary school | 495 | 7.1 |  | 258 | 61.6 |  | 753 | 25.8 |  |
| Intermediate school | 651 | 11.7 |  | 357 | 67.5 |  | 1,008 | 31.4 |  |
| Secondary school | 2,675 | 13.9 |  | 595 | 78.8 |  | 3,270 | 25.7 |  |
| Higher education | 1,478 | 15.2 |  | 366 | 89.9 |  | 1,844 | 30 |  |
| **Monthly income** |  |  |  |  |  |  |  |  |  |
| <5000 | 1,433 | 7 | <0.001 | 1,147 | 73.4 | <0.001 | 2,580 | 36.5 | <0.001 |
| 5000-<10000 | 2,415 | 10.3 |  | 486 | 79.2 |  | 2,901 | 21.9 |  |
| 10000-<15000 | 1,280 | 14.1 |  | 136 | 68.4 |  | 1,416 | 19.3 |  |
| 15000-<20000 | 612 | 21.6 |  | 69 | 50.7 |  | 681 | 24.5 |  |
| 20000+ | 611 | 17.5 |  | 87 | 54 |  | 698 | 22.1 |  |
| **Chronic disease** |  |  |  |  |  |  |  |  |  |
| Yes | 1,830 | 12.7 | 0.32 | 278 | 68 | 0.05 | 2,108 | 20 | <0.001 |
| No | 4,521 | 11.8 |  | 1,647 | 73.6 |  | 6,168 | 24.2 |  |
| **Working Status** |  |  |  |  |  |  |  |  |  |
| Employed | 2,507 | 14.5 |  | 1,462 | 73.9 |  | 3,969 | 36.4 |  |
| Unemployed | 999 | 8.2 | <0.001 | 68 | 73.5 | 0.136 | 1,067 | 12.4 | <0.001 |
| Housewife | 2,104 | 10.4 |  | 389 | 68.9 |  | 2,493 | 19.6 |  |
| Retired | 741 | 13.9 |  | 6 | 50 |  | 747 | 14.2 |  |
| **Marital status** |  |  |  |  |  |  |  |  |  |
| Married | 3948 | 13.9 |  | 1413 | 76.6 |  | 5361 | 30.4 |  |
| Never married | 1986 | 9.8 | <0.001 | 470 | 64.5 | <0.001 | 2456 | 18.5 | <0.001 |
| Divorced &<br>Widowed | 417 | 6 |  | 42 | 40.5 |  | 459 | 9.2 |  |
| **Self-rating of**<br>**health status:** |  |  |  |  |  |  |  |  |  |
| Very Good | 4,352 | 12.2 | <0.001 | 1,579 | 72.9 | 0.002 | 5,931 | 28.4 |  |
| Good | 1,219 | 12.3 |  | 285 | 76.1 |  | 1,504 | 24.4 | <0.001 |
| Fair | 596 | 13.4 |  | 46 | 63 |  | 642 | 17 |  |
| Bad | 182 | 3.3 |  | 15 | 33.3 |  | 197 | 5.6 |  |
| **Nationality** |  |  |  |  |  |  |  |  |  |
| Saudi | - | - |  | - | - |  | 6351 | 12.1 | <0.001 |
| Non-Saudi | - | - |  | - | - |  | 1925 | 72.8 |  |

\*Based on chi-squared test

The average number of health care visits during the past 12 months by nationality and other characteristics are shown in Table 3. The mean number of visits was significantly higher among Saudis (mean±SD; 1.73±1.351) than non-Saudi (0.92±1.139). The mean value was higher in females (mean±SD; 1.71±1.349) compared to males. In both Saudi and non-Saudi populations, the elderly group (<60 years) had higher average health facility visits compared to adult (26-59 years) and (18–25) groups. The mean values were significantly higher among lower education level in Saudis and non-Saudis (mean±SD; 1.64±1.336) than the higher education level (mean±SD; 1.54±1.336). In both Saudis and non-Saudis, the mean values of the number of visits were significantly higher among those with high monthly income (+20,000 SR) (mean±SD; 1.96±1.453) compared to low-income groups. This were significantly higher among the divorced & widowed (mean±SD; 1.83±1.310) than the married and never married groups. The values were found to be higher among those who had no insurance (mean±SD; 1.69±1.368) and those who had chronic disease (mean±SD; 1.92±1.346), which applies to both Saudis and non-Saudis. The average number of visits were significantly lower for those who perceived their general health as being very good (mean±SD; 1.40±1.325) compared to those who rated their health as bad (mean±SD; 2.50±1.231). We found similar differences in the number of visits according to the perceived health status group among Saudi and non-Saudi populations.

**Table 3.**
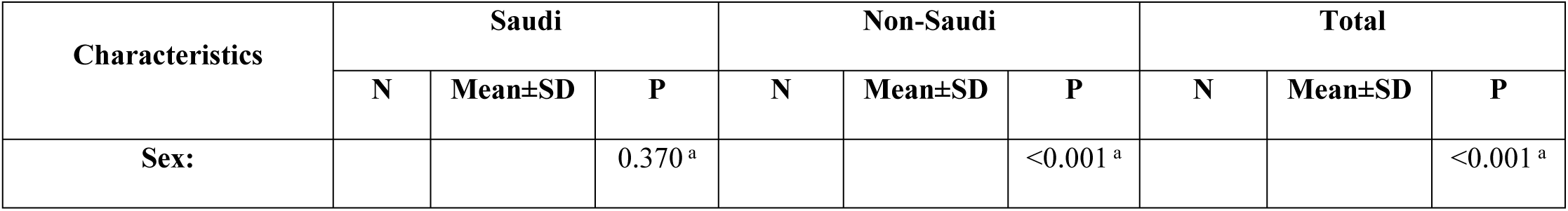
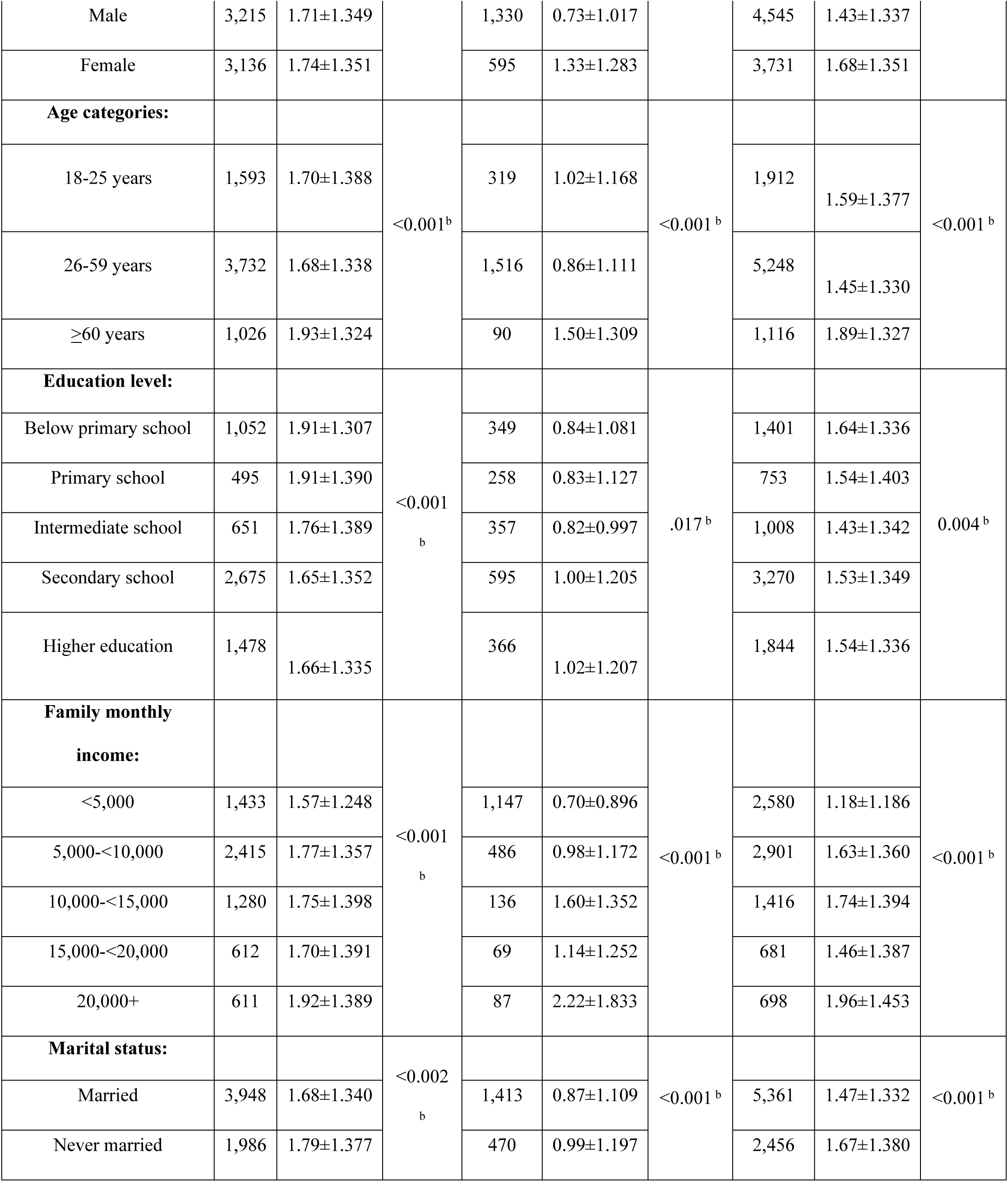
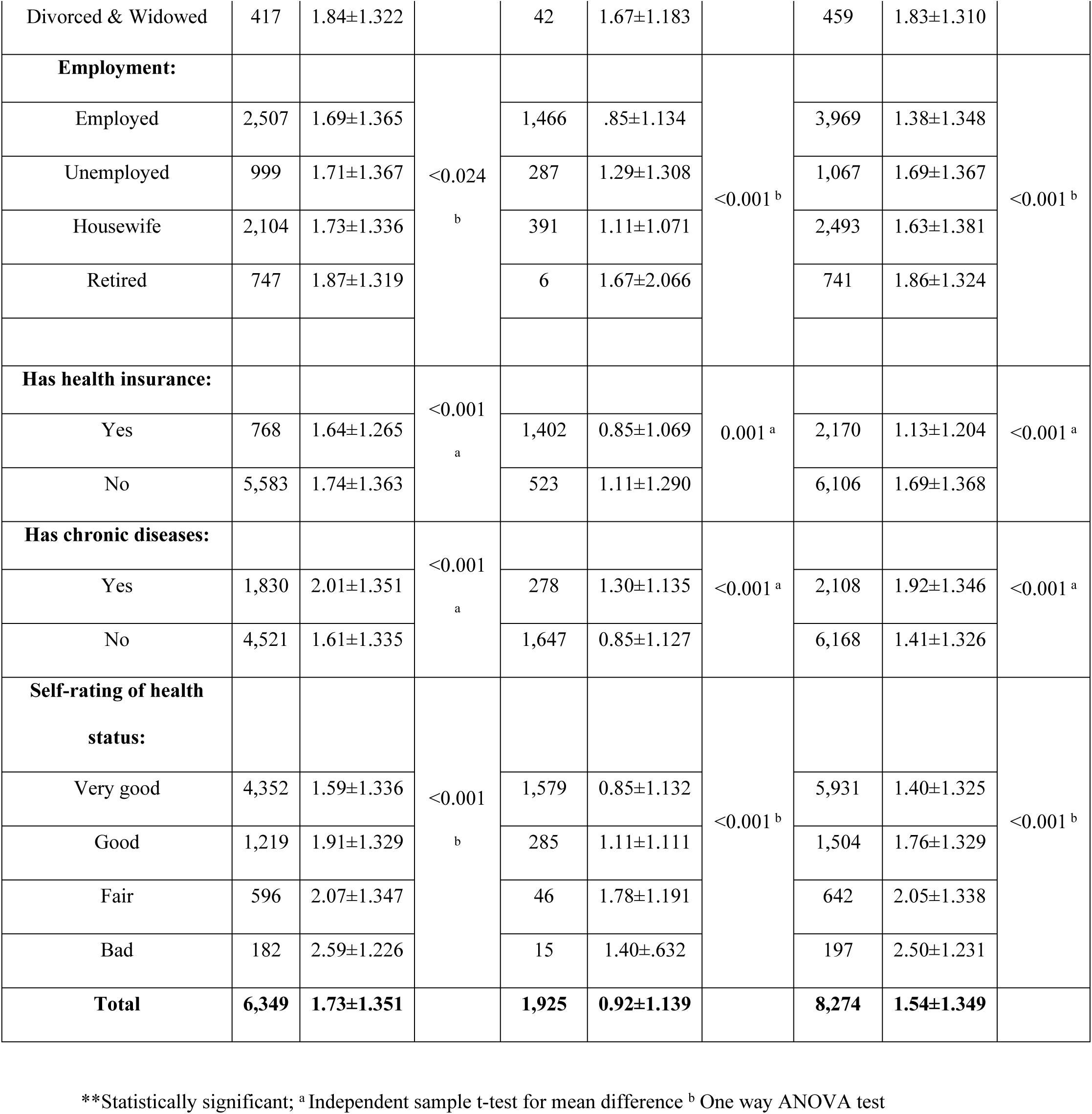
Average number of health facility visits for outpatient care during the past 12 months by respondents’ characteristics.

The regression analysis results examining the factors influencing the enrollment of health insurance are presented in Table 4. The likelihood to be enrolled in insurance was 27 folds in non-Saudis (OR=27.127; 95% CI=22.985-32.017) compared to Saudis. Males were found to be more likely to be insured by two folds (OR=2.090; 95% CI=1.700-2.569). The higher-income individuals (15,000-20,000 SR) were two times (OR=1.860; 95% CI, 1.453-2.380) more likely to enroll in the insurance scheme compared to low-income individuals (<5,000 SR). Also, when compared to those with below primary school education level; the odds to be insured was almost four folds in those with higher education (undergraduate and above) level (OR=3.584; 95% CI, 2.815-4.563). The likelihood of those who perceive health as being good was two folds (OR=2.767; 95% CI, 1.350-5.668) more likely to have insurance compared to those who perceive health as bad. We found that being never married decreased the odds to be enrolled in insurance compared to being married respectively.

**Table 4.** Predictors of the health insurance enrollment among the study group.

| Variables | Description | Odds ratio (95% CI) |
| --- | --- | --- |
| Constant |  | 0.023*** (0.010; 0.054) |
| Chronic disease | No (Ref) | 1 |
|  | Yes | 1.068 (0.882; 1.292) |
| Perceived the Health status | Bad (Ref) | 1 |
|  | Fair | 4.141*** (1.994; 8.602) |
|  | Good | 2.767** (1.350; 5.668) |
|  | Very good | 2.343* (1.994; 8.602) |
| Nationality | Saudi (Ref) | 1 |
|  | Non-Saudi | 27.127*** (22.985; 32.017) |
| Sex | Female (Ref) | 1 |
|  | Male | 2.090*** (1.700; 2.569) |
| Monthly income | <5000 (Ref) | 1 |
|  | 5000-<10000 | 1.043 (0.881;1.235) |
|  | 10000-<15000 | 1.166 (0.945;1.438) |
|  | 15000-<20000 | 1.860*** (1.453;2.380) |
|  | 20000+ | 1.426** (1.104;1.841) |
| Education level | Below primary school (Ref) | 1 |
|  | Primary school | 1.154 (0.882;1.509) |
|  | Intermediate school | 1.810*** (1.408;2.327) |
|  | Secondary school | 3.197*** (2.553;4.002) |
|  | Higher education (undergraduate and above) | 3.584*** (2.815;4.563) |
| Age | 18-25 years (Ref) | 1 |
|  | 26- 59 years | 0.933 (0.705;1.236) |
|  | >60 years | 0.815 (0.571;1.163) |
| Marital status | Married (Ref) | 1 |
|  | Never married | 0.653*** (0.541;0.787) |
|  | Divorced & Widowed | 0.513*** (0.345;0.762) |
| Log likelihood |  | -1353.00 |
| Chi-squared (DF) |  | 2,199 (1,444) |
| N |  | **8,274** |

Note: \*\*\* p<0.01; \*\* p<0.05; \* P<0.10

The regression analysis findings on the factors of the utilization of health services are presented in Table 5. We found health insurance enrollment was negatively associated (Coefficient = -0.107, P<0.01) with the utilization of outpatient health services. Insured individuals had lower utilization compared to uninsured individuals. The male population had lower utilization of services compared to the female population.

**Table 5.**
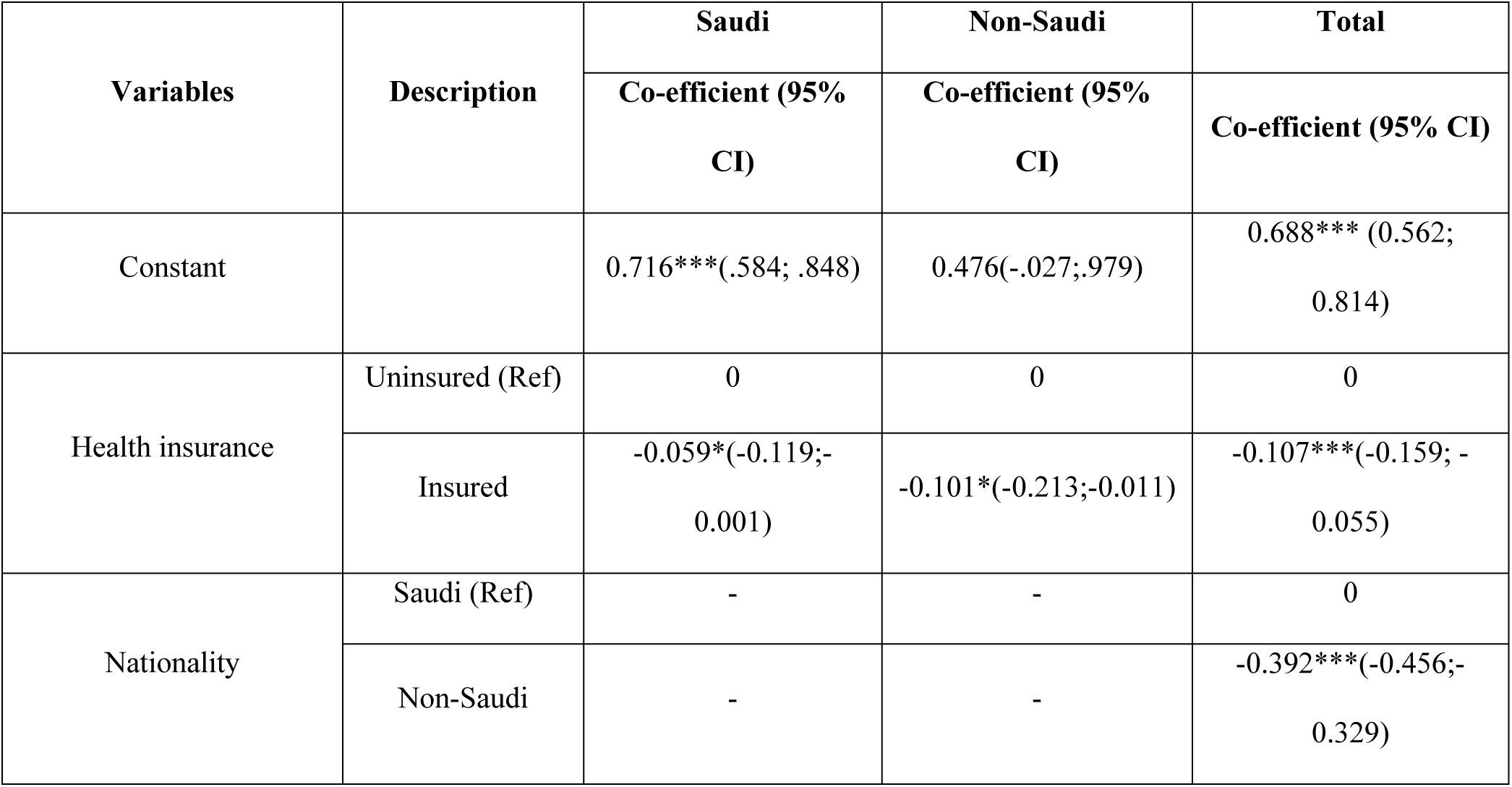
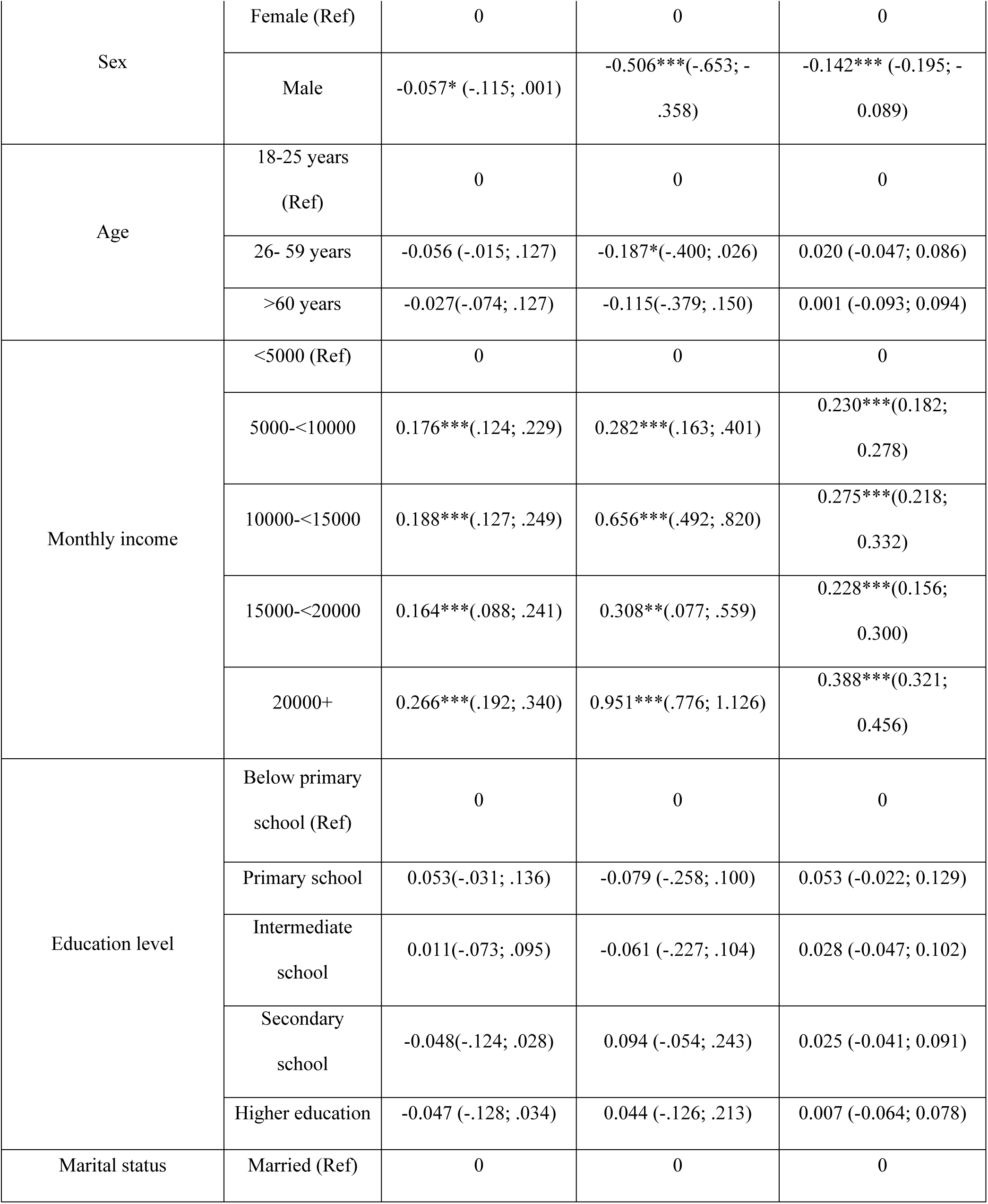
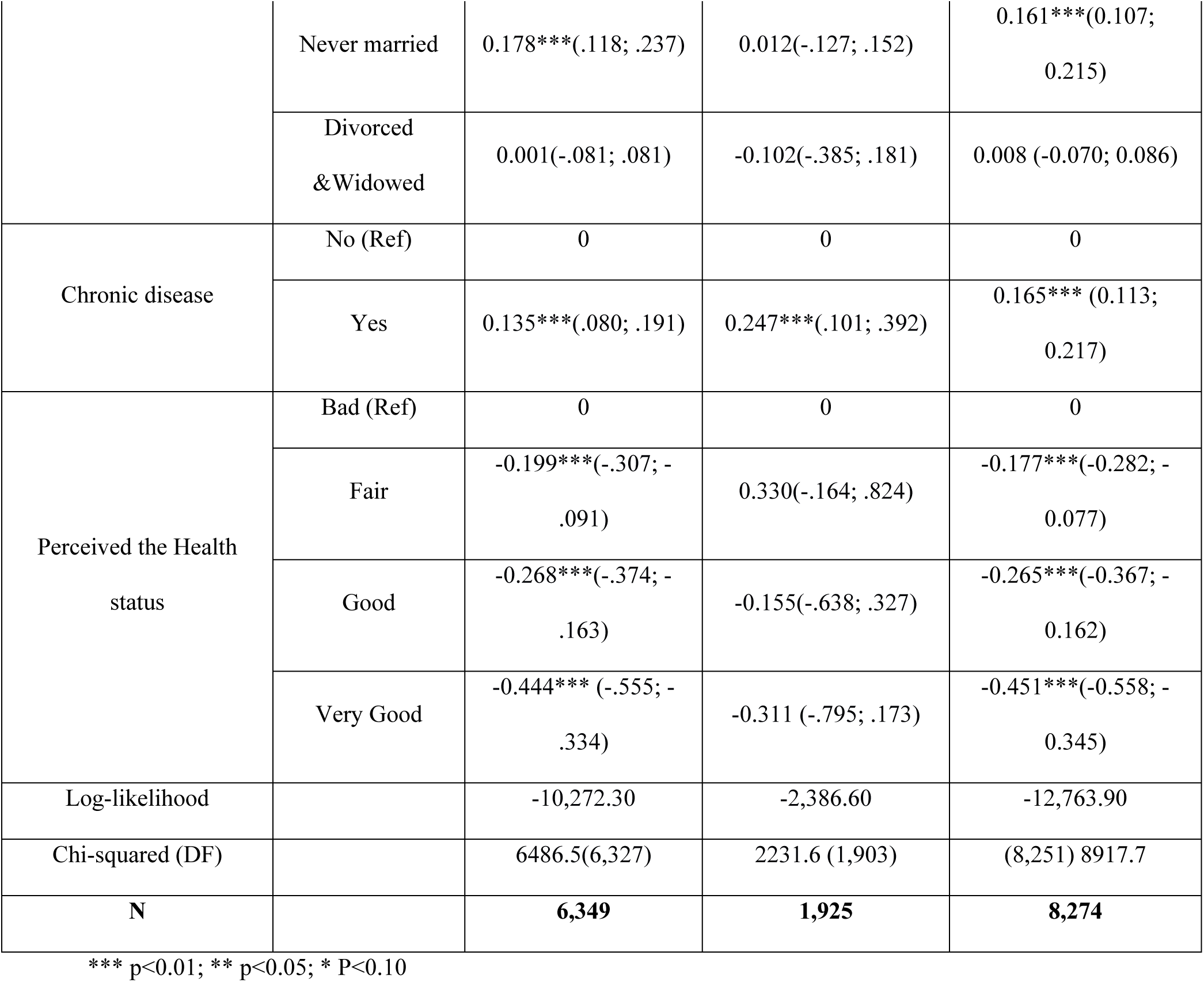
Predictors of the utilization of outpatient care among the study group.

If we compared those individuals with a lower monthly income (<5,000 SR), the utilization of health services was higher among individuals with higher monthly income (20,000 SR or more) (coefficient = 0.388 95% CI= 0.321; 0.456). It was found that never-married individuals have higher health care services utilization (Coefficient = 0.161 95% CI= 0.107; 0.215) compared to married individuals. People suffering from chronic conditions had higher utilization of health services compared to people who had no such conditions.

We conducted separate regression models for Saudi and non-Saudi populations to further examine the association between health insurance enrollment and utilization of services among these two distinct populations. In both population groups, health insurance enrollment was associated with lower utilization of health services.

## DISCUSSION

The study highlighted the determinants of health enrollment and the association of health insurance status with the utilization of outpatient health care services in Saudi Arabia. We found that non-Saudi nationality, higher monthly income, male sex, married marital status, higher education level and good perceived health status led to health insurance enrollment. Further, the factors that increased the utilization of outpatient services were being Saudi, female, with high monthly income, never married, having chronic diseases and perception of bad health status. We found respondents having health insurance were less likely (OR=-0.107) to utilize outpatient services compared to those who don’t have insurance.

### Determinants of health insurance enrollment

The study showed that there was a significantly higher proportion of non-Saudis who were covered by health insurance. This is an impact of the implementation of the first phase of the CHI scheme that necessitated compulsory insurance for all those who are working in the private sector where mostly expatriates are employed (16). In Saudi Arabia, about 80% of the workforce in the private sector are non-Saudi expatriates and the rest are Saudi citizen (27). Al-Hanawi and colleagues found in 2016 that in Saudi Arabia 38% of the total population are covered under CHI scheme (19). This includes approximately 2.7 million Saudi citizens, which represents about 22% of the insured population, and around 9.4 million expatriate residents (non-Saudis), accounting for about 78% of the insured population. The remaining Saudi citizens have free access to publicly funded health care facilities and can use the private health care facilities on a fee for services basis at the point of use or can purchase private insurance (19). While Saudi citizens readily have free access to publicly funded health care services, some opt for the uptake of private insurance due to potential advantages such as avoiding relatively long waiting times and access to a wider range of services, including luxurious services like more appointment options, extra accommodation services, and additional health services (28,29). Moreover, the GASTAT showed 21.3% prefer to visit private health care services by cash payment for the same reasons (17)

Age was found to be a significant predictor of health insurance uptake for Saudis and non-Saudi; with an increased percentage among those who were aged 26-59 years (13.6%) compared to those aged 18-25 years (9.5%) while non-Saudi 26-59 years (75.2%) compared to aged >60 years (58.9%) (Table 2). This may be because the implementation of the first phase of insurance was compulsory for employees which includes only the working-age population (30). which accords with what had been found in a study conducted in Saudi Arabia which reported that those aged 19–59 year olds were willing to pay more for National Health Insurance compared to participants aged 60 or older which indicates that young individuals were more likely to pay more money for NHI compared to older ones; that has been attributed to that younger people might have fewer financial responsibilities than older ones (29)

The individuals with higher income were two times (OR=1.860) more likely to enroll in the insurance scheme compared to low-income individuals, which accords with the findings of multi-country research in Nigeria, South Africa, and the United States of America (31,32). This may be because low-income self-employed Saudi citizens are less likely to have health insurance due to the employer-based insurance system. For this group of population, the health insurance is not mandatory, and the willingness to enroll or pay can be greatly influenced by individual monthly income (25). Those with lower incomes may be more likely to refrain from enrolling in health insurance due to their financial condition. Therefore, it is important to consider the affordability of insurance when designing the National Health Insurance Program (33). The non-Saudi individuals required a health insurance irrespective of their employment condition (e.g., employed, self-employed or unemployed) to legally stay in the country.

Males were more likely to have health insurance than females, which is in the of the previous study in Saudi Arabia (34). This result may be attributed to the workforce composition in Saudi Arabia (79.9% of males working compared to 21.1% of females), which is characterized by male predominance (35) (36). Moreover, considering that employed individuals are compelled to obtain insurance, this could elucidate the heightened prevalence of insurance coverage among males. In contrast, in most European countries as Germany where the gender gap in health insurance coverage is less pronounced due to the nature of the universal health system implementation as health insurance is mandatory for all residents regardless of employment status.(37)

The study revealed that individuals who were employed exhibited a higher propensity to possess health insurance, which comes in line with the implementation of the initial phases of the cooperative health insurance program in Saudi Arabia. These initial stages entailed obligatory enrollment in insurance plans for employed individuals in the private sector (4). The study showed that the married were more likely to have health insurance, which comes in congruence with the previous study which revealed that being married was associated with a tenfold increase in the likelihood of owning health insurance compared to patients who were never married, which suggests that marriage may encourage insurance ownership due to factors like the desire to protect children and mitigate the risk of catastrophic health care expenses (38).

The level of education has been identified as a determining factor of the enrollment of health insurance. This finding aligns with the results of a previous study conducted in Ghana (39) and low-and middle-income countries (40). It is worth noting that individuals with higher levels of education tend to possess a better understanding of the insurance policy compared to those with lower educational backgrounds. Additionally, individuals with higher educational backgrounds have the opportunity to comprehend the fundamental concepts and advantages of the health insurance scheme (41). Moreover, individuals with higher educational background become more knowledgeable about activities that directly impact the health and well-being of their families (39). Since higher educated individuals more likely to have employed compared to individual with low educational qualification, they have more chance to enroll health insurance through their employment. The high percentage of those who perceived that their health is good among the insured, could be attributed partly to the findings in the current study that the majority of insured are expatriates, and it is known that expatriates in Saudi Arabia are mandated to undergo pre-employment medical examinations before they enter the country to ensure their fitness for the job (42)

### Determinants of utilization of outpatient

The study demonstrated that the insured were less likely to utilize health outpatient services, which aligns with a previous systematic review by Zhang et al. (2020). This finding contradicts the expectation that insurance would enhance access to health care services(43). One possible explanation is that the utilization of medical services is more directly correlated with the need for the services rather than with insurance coverage (44). The majority of the insured individuals in our study were non-Saudi (expatriates), who are commonly younger (with an ages range between 30-39 years) compared to the entire population (median age of 35.5 years) (13). These individuals exhibit a relatively low prevalence of chronic ailments and tend to perceive their health as being in good or very good condition (52.8%) resulting in a relatively lower need for health services. Other contributing factor, includes the limited understanding of the insurance policy (health care literacy), may help elucidate the reasons behind the limited utilization of health care services by the insured individuals (13,45). Another explanation has been addressed by AlNemer (2018) that the expatriates perceive that the policy of insurance they possess has a weak quality with limited benefits, as it is often done by the employer with the lowest premium. On the other side, the relatively high utilization by Saudis could be understood from the fact that have free access to health services (46). It should be noted here that we included only outpatient service utilization in our analysis and including all types of services (e.g., primary, inpatient) may provide different results.

Higher monthly income was found to be associated with increased utilization of health services. Al-Hanawi et.al (2021) and Ukert B et.al (2022) refers the positive relationship to the highest rate of utilization of health services among those with high-income (47,48). It had been assumed that “those with better incomes may be in a position to support themselves to use private health care services or purchase health insurance, which has been found to contribute to easy access to the health care” (12).

Older age has been associated with higher utilization of health care services (Table 3). Elderly people have a higher prevalence of non-communicable diseases which eventually leads to higher utilization of health care services (49). Females are more likely to utilize health care services, which accords to past research (50). However, the association between higher female morbidity and increased health care utilization remains inconsistent (51). Notably, women reported significantly poorer self-perceived health status, which may partially explain their greater use of certain services like general practitioner visits and diagnostic procedures (52). The study of Obuchowska et al (2020) refers to women who visit gynecologists frequently, younger women often visit for check-ups for pregnancy tests or contraceptive needs and preventive care, while reasons for visits include addressing specific health concerns such as pelvic pain or abnormal bleeding (53). The never-married individuals demonstrated a higher frequency of outpatient health care service utilization. Never married tend to access health care services more frequently than married, such as visits to general practitioners, psychiatrists, and psychologists; divorced in general, also report a higher prevalence of unmet health care needs, which is often associated with higher levels of depressive symptoms (54). Moreover, large proportion of single females 73.6 % tend to undergo cosmetic surgery to improve their confidence (55).

Having chronic disease was associated with a significant increase in utilization of health services, as chronically ill patients need frequent visits for regular check-ups and receive treatment, and they are more prone to other complications (56,57). In Saudi Arabia, the reports pointed to the relatively high prevalence of preventable chronic diseases such as hypertension, diabetes, and hypercholesterolemia (58). In a study conducted by Alsubaie et al. in Saudi Arabia, it had been shown that chronic disease doubles the likelihood of utilization of health care services (OR = 2.02) (59). The results of the current study showed that the perceived health condition was a substantial determinant for the utilization of health services with a significantly higher number of visits from those who ranked their health as being “bad”, which is supported by the theoretical model of health utilization, indicating that the perceived needs for health care is a main predictor of utilization of health services (60,61).

## STRENGTHS AND LIMITATIONS

The main strength of the study is based on nationwide data including a representative sample from all geographic regions of Saudi Arabia. Further strength could be the application of multiple regression models in this study for identifying possible determinants of insurance enrollment and association between health insurance enrollment and health care utilization while adjusting for confounders. However, due to cross sectional nature of the FHS, one inherited limitation in that this study is unable to conclude any causal relationships. Another limitation of the study is the recall bias when reporting the health care utilization (1 year) and other information by the respondents. We were able to access only FHS 2018 data for this study while a recent version of the dataset is released by the General Authority for Statistics after completion of the analysis. We were unable to include inpatient care utilization in our analysis due to the unavailability of this information in the dataset we received from the GASTAT. These limitations should be considered while interpreting and using the findings of this study.

## CONCLUSIONS

The study investigated the determinants of health insurance enrollment and the utilization of health care services in Saudi Arabia and found that non-Saudi, males, having high income, and higher level of education, and perceived good health status are more likely to be enrolled in health insurance. Factors increasing utilization of outpatient services were being female, having a high monthly income, being never married, having chronic diseases, and the perception of bad health. However, health insurance was associated with lower utilization of outpatient services. Consequently, policymakers should take more targeted measures that address the challenges and opportunities associated with health insurance enrollment and health care service utilization. Introducing targeted interventions oriented toward improving health insurance literacy and understanding insurance benefits for the insured population could be helpful to improve the current level of health service utilization. These measures could involve creating a robust education campaign and enhancing health services accessibility to diverse population groups. To guarantee every resident has equitable access to health care, we strongly suggest the development of a health insurance program based on a different financing solution. Future research can be aimed at proving the long-term effectiveness of health insurance in ensuring diversified types of health service (e.g. including inpatient and primary health care) and impact on quality, and health outcomes for whole populations.

## Data Availability

Data cannot be shared publicly because the data were obtained from the Saudi Statistical Authority, which requires researchers to outline the study's objectives and specify the data needed. Data are available upon request from the Saudi Statistical Authority. Here is the

## DECLARATIONS

### Availability of data and materials

The access to the datasets generated and/or analyzed during the current study can be obtained through the General Authority for Statistics in Saudi Arabia, as they are not publicly available due to privacy, confidentiality, and other restrictions.

### Fund

The authors have not received any funding or benefits to conduct this study.

### Authors’ contributions

Conceptualization: Khaled Shaeel Althabaiti, Jahangir Khan, Sayem Ahmed, Monica Hunsberger

Methodology: Khaled Shaeel Althabaiti, Jahangir Khan, Sayem Ahmed, Monica Hunsberger

Data curation: Khaled Shaeel Althabaiti

Validation: Khaled Shaeel Althabaiti

Formal analysis: Khaled Shaeel Althabaiti, Sayem Ahmed, Jahangir Khan, Monica Hunsberger Writing - original draft: Khaled Shaeel Althabaiti, Sayem Ahmed, Monica Hunsberger, Jahangir Khan

Writing – review and editing: Khaled Shaeel Althabaiti, Sayem Ahmed, Monica Hunsberger, Jahangir Khan

## Acknowledgement

Each author has played a role in completing this study. We are grateful to the General Authority for Statistics (Saudi Arabia) for providing us the FHS 2018 dataset for conducting this analysis.

## List of abbreviations

KSA: Kingdom of Saudi Arabia
FHS: Family Health Survey
MOH: Ministry of Health
WHO: World Health Organization
GCC: Gulf Cooperation Council
GASTAT: General Authority for Statistics

